# The Physical Exercise and Cardiovascular Adaptation Monitoring in Pregnancy (PE-CAMP) Randomised Controlled Trial Protocol

**DOI:** 10.1101/2022.04.27.22274359

**Authors:** O. Roldan-Reoyo, K A. Mackintosh, M A. McNarry, S. Jones, S. Emery, O. Uzun, M J. Lewis

## Abstract

**Background:** Exercise can be beneficial to cardiovascular system function, but its influence during pregnancy is less well understood. Heterogeneity in research studies has led to a lack of consensus on whether physical exercise during pregnancy can elicit cardiovascular adaptations during a period in which the cardiovascular system is already being challenged. Furthermore, little research has been conducted regarding the influence of physical exercise on foetal heart rate (FHR). This paper presents the protocol for a randomised controlled trial which will evaluate the influence of supervised antenatal physical exercise on cardiovascular adaptations during and after pregnancy, as well as the FHR response to acute and chronic maternal exercise.

**Methods:** The PE-CAMP Study (Physical Exercise and Cardiovascular Adaptation Monitoring in Pregnancy) is a randomised controlled trial (RCT) in which pregnant women will be randomised into an intervention group (INT), which attended supervised physical exercise programme up to three days per week, or a comparison group (COMP) which followed standard health care. All participants were assessed at three time-points i) 18-22 weeks pregnant, time-point 1; ii) 33-37 weeks pregnant, time-point 2; and iii) 12-16 weeks postnatal, time-point 3. A standardised experimental protocol was used for data collection, including body composition assessment, upper-body flexibility and strength assessment, physical activity assessment via questionnaires and acceletometry, and haemodynamic and cardiovascular evaluation before, during and after an acute 10-minute exercise bout. Foetal heart rate will be assessed at the time-point 2 before, during and after acute exercise.

**Discussion:** Although it is necessary and informative to continue investigating the effects of exercise on maternal cardiac and haemodynamic responses using specific laboratory-based tests, it is also critical to evaluate these influences during activities that are more achievable and realistic for pregnant women. The PE-CAMP study will provide data on the cardiac and haemodynamic responses to a typical acute bout of exercise, which could help inform future decisions and policies on maternal exercise prescription made by maternity healthcare providers and exercise professionals.

**Clinical Trials Registration Number:** NCT03748888

## 1. Introduction

Cardiovascular disease (CVD) is the leading cause of death worldwide and a significant cause of mortality during pregnancy (1–4). Indeed, in the United Kingdom (UK), the maternal mortality rate from cardiac disease has remained unchanged since 2003, accounting for 56% of all indirect maternal deaths (1).

It is widely known that healthy pregnancy elicits adaptations in maternal haemodynamics via blood volume expansion, increased heart rate (HR), stroke volume (SV) and cardiac output 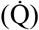 and decreased blood pressure (BP) and total peripheral resistance (TPR) (5–9). Furthermore, pregnancy induces a progressive change in the cardiac autonomic nervous system (ANS), with sympathetic nervous system activation and parasympathetic nervous system withdrawal (5,10– 12), consequently affecting maternal heart rate variability (HRV; 13,14). The majority of cardiac parameters influenced by pregnancy return to pre-pregnancy values postnatally (8,15). However, some studies have reported a persistently elevated SV and end-diastolic volume at both six and 12 weeks postnatal (16), and significantly increased sympathetic regulation six weeks postnatal compared to the end of the first trimester (17). This has led to suggestions that pregnancy-induced cardiovascular changes might persist later in life and could influence subsequent pregnancies (18,19).

Despite numerous previous studies focusing on cardiovascular adaptations and cardiac function during pregnancy, more research is needed, especially with regard to how exercise affects pregnancy (20). Exercise can be beneficial to cardiovascular system function and CVD prevention in non-pregnant populations through an enhanced ANS function and improved left ventricular function, both of which lead to a higher 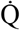 and 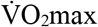 (21,22), but its influence during pregnancy is less well understood.

International physical activity (PA) recommendations for pregnant women agree on the potential benefits of being physically active during pregnancy (23–26). PA can help to avoid excessive gestational weight gain and reducing the risk of pregnancy-related complications, such as hypertensive disorders, gestational diabetes and the risks of having a caesarean section delivery (27–30). Contrary, not meeting PA recommendations during pregnancy can trigger co-morbidities, such as being overweight or obese, which are associated with pregnancy-related diseases (pre-eclampsia, gestational diabetes, higher risk of c-section delivery, foetal macrosomia; 31–34). These complications can influence physiological function, including cardiovascular and ANS function. Therefore, such variables represent important potential markers of appropriate physiological adaptations to pregnancy.

Previous studies investigating the effects of PA and exercise on the maternal cardiovascular system have used heterogenous methods and protocols leading to inconsistent results, potentially masking the effects of prenatal exercise. May et al. (35) reported that active pregnant women had a significantly lower HR and higher HRV compared to sedentary pregnant women, especially at 28 weeks pregnant. Carpenter et al. (35, 36) investigated the haemodynamic and cardiac responses to an exercise intervention at three time points during pregnancy (12-16, 26-28, and 34-36 weeks pregnant. Specifically in comparison to control group, chronic exposure to exercise during gestation was shown to lead to an altered cardiovascular response following a supervised exercise intervention. These differences were statistically significant in late gestation, showing greater reductions in the root mean squared of successive differences between normal beats (RMSSD) and standard deviations of R-R intervals (SDRR), and increased QT variability in the supine position in the intervention group (36). Furthermore, whilst only diastolic BP during exercise was lower in the intervention group during the second trimester, postnatally the intervention group displayed a greater SV and end-diastolic index (EDI) and a lower TPR and diastolic BP during exercise (35). In contrast, Baciuk et al.(37) found no differences across three time points during pregnancy (18-20, 22-26, 32-26 weeks pregnant) in cardiovascular and cardiorespiratory variables (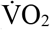 uptake, BP and HR) following a swimming-based intervention. Similarly, Perales et al.(38) reported no significant effect of supervised exercise three times per week on cardiac function analysed via echocardiography at two different points during pregnancy (20 and 34 weeks pregnant). However, it is pertinent to note that participants were assessed at rest which may fail to reveal important changes in the dynamic response of the cardiovascular system. There is therefore a lack of consensus regarding whether exercise interventions during pregnancy can elicit cardiovascular adaptations during a period in which the cardiovascular system is already being challenged.

Foetal ANS function during pregnancy and early childhood is key for postnatal development and progression to a healthy adulthood (39–43). However, little research has considered the influence of chronic exercise exposure on the foetal heart rate (FHR) or cardiac ANS function. May et al. (44) assessed FHR and foetal HRV during maternal rest via magnetocardiography at different time points during pregnancy, reporting that foetuses from physically active pregnant women had a higher HRV and lower FHR at 36 weeks pregnant. To date, this is the only study showing significant differences in foetal HRV active vs non-active pregnant women. In contrast, a recent systematic review and meta-analysis focusing on FHR and including 91 studies concluded that there is no clinically relevant response of FHR to either acute or chronic maternal exercise (45). These contradictory findings may be the result of different protocols, such as sample characteristics, sample size, assessment protocol, equipment used to evaluate, exercise test, and assessment of PA levels (45).

The present protocol therefore sought to generate the first combined evidence regarding the impact of a supervised antenatal physical exercise programme designed for pregnant women following national and international PA guidelines on the maternal cardiovascular system and on the cardiac adaptation of the foetus. The study will propose and use a standardised protocol to assess maternal cardiac and haemodynamic function during and after pregnancy and will compare the foetal cardiac response to acute maternal exercise in women who were randomly assigned to a physical exercise intervention group (INT) or usual health care comparison group (COMP).

The main objectives of the study are: i) to longitudinally assess maternal cardiac and haemodynamic responses to quantified antenatal physical exercise and physical activity during and after pregnancy; ii) to analyse the influence of the antenatal physical exercise programme on other physical fitness components (upper-body flexibility and strength, body composition) during and after pregnancy; iii) to assess the influence of the antenatal physical exercise programme on the FHR response to acute maternal exercise; and iv) to evaluate the influence of the exercise intervention on pregnancy outcomes and other quality of life-related variables (quality of life, sleep quality and urinary incontinence).

We hypothesise that women randomised to a supervised bespoke physical exercise programme will show improved cardiac function, as reflected in a reduced HR and increased HRV, SV and 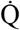 and that this will be sustained in the postnatal period. Moreover, we hypothesise that foetuses from women randomised to the antenatal physical exercise programme will show lower FHR.

## 2. Methods

### 2.1. Study design

The Physical Exercise and Cardiovascular Adaptation Monitoring in Pregnancy (PE-CAMP) Study was a randomised controlled trial (RCT) (registration number: NCT03748888) designed to investigate the influence of supervised exercise on maternal cardiovascular adaptations to pregnancy and the foetal cardiac response to maternal chronic and acute exercise. The PE-CAMP study was a multicentre trial based in South Wales, UK and conducted in collaboration between academia and a local health board from April 2018 to October 2020. Eighty-eight pregnant women were sought to be randomised on a 1:1 basis to two different groups, INT and COMP. The INT was based on a supervised physical exercise programme designed for pregnant women involving three sessions per week and starting no later than 16 weeks pregnant and proceeding until 36 weeks pregnant. The COMP did not receive any intervention and followed standard health care. Participants were assessed at three timepoints during pregnancy (18-22 weeks pregnant, time-point 1; 33-37 weeks pregnant, time-point 2; and 12-16 weeks postnatal, time-point 3; Figure 1). Ethics approval was obtained from the corresponding National Health System Research Ethics Committee (South West Wales, UK; REC Reference: 17/WA/0414; Protocol Reference: RIO 027-17. Protocol version 5: 18^th^ December 2018). All study procedures followed the ethical principles of the Declaration of Helsinki under its last update in October 2013 (46). Monitoring of the study was done accordingly to the Standard Operating Procedures published by the Health Research Authority UK https://www.hra.nhs.uk/about-us/committees-and-services/res-and-recs/research-ethics-committee-standard-operating-procedures/. This guidance was also used to inform the relevant Ethics Committee of protocol amendments. Moreover, the protocol is being reported under the SPIRIT 2013 Statement for clinical trials (ESM1; 47) and the CERT statement (ESM2; 48). To ensure that the study was conducted appropriately, an international steering committee was created.

**Figure 1.**
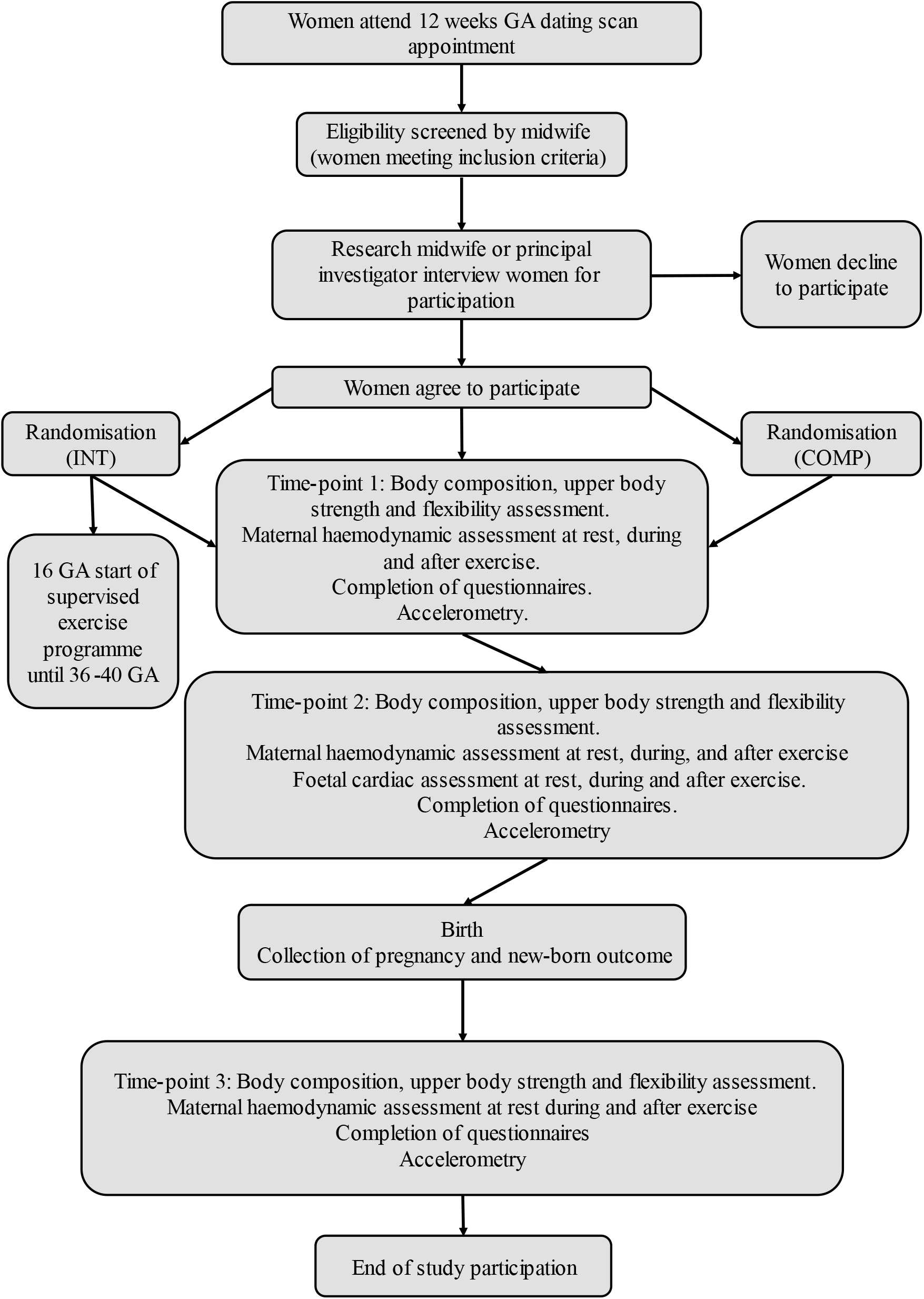
Flow chart of study participation.

To increase study participation and promote social engagement with the study, the research team created the Being Active Benefits You (BABY) group. The BABY group consisted of monthly meetings where all study participants irrespective of randomisation were invited to attend informal talks and workshops focused on topics such as pelvic floor health, abdominal diastasis prevention and treatment, mental health during and after pregnancy, early infant stimulation, and birth plan writing.

### 2.2. Sample size

The original sample size calculation was based on the study of Perales (50). Specifically, to obtain a minimum difference in SV of 5.08 ml with a standard deviation of 12.79 ml, a power of 80% and a confidence of 95%, 34 participants were required per group. Assuming a 30% loss to follow-up per group (51), the final target sample size was a minimum of 44 participants per group.

### 2.3. Recruitment

There were two ways of recruitment. The first one was done at a clinical setting where pregnant women attending their twelve-week dating scan appointment were screened by a midwife, to assess them for eligibility for participation, based on the stated inclusion and exclusion criteria (Table 1). Afterwards, eligible participants were approached by the PI or a research midwife to discuss their possible participation in the study. The second way of recruitment was done via posters advertising the study, which were placed in maternity units, general practitioner surgeries and sports centres. Pregnant women expressing an interest in participation from reading an advertising poster needed then to be screened by their midwife or main healthcare provider in order to check study eligibility. Those willing to participate were required to sign an informed consent form (ESM 3) after reading the participant information sheet and had the opportunity to speak to the PI if they had questions relating to the study. Participants who developed any medical/obstetric complications considered a contraindication to any form of physical activity were removed from the study.

**Table 1.** Inclusion and exclusion criteria.

| Inclusion criteria | Exclusion criteria |
| --- | --- |
| Women with healthy, low risk pregnancy | Known complications of pregnancy |
| Age 18-40 years | Known contraindications to physical exercise |
| Single pregnancy | Known major cardiovascular or respiratory pathology |
| Pregnancy stage $\leq 16$ weeks at the point of consent | Three or more previous term pregnancies |
| Two or fewer previous term pregnancies (as physiology can be altered by multiple pregnancies) | Inability to communicate in English or Welsh |

### 2.4. Randomisation

Participants were randomised on a 1:1 basis to either the INT or COMP following a stratified and block randomisation procedure (52) based on pre-pregnancy BMI (BMI < or > 25 kg•m^−2^) and pre-pregnancy PA levels (inactive = < 150 min • week^-−2^, active = > 150 min • week^-−2^; based on the UK recommendations for PA for adults; 26). Therefore, participants were evenly allocated across four groups (Active, BMI < 25 kg•m^−2^; Active, BMI > 25 kg•m^−2^; Inactive, BMI < 25 kg•m^−2^; and Inactive, BMI > 25 kg•m^−2^). Due to the characteristics of the study, participant’s group allocation was only blinded to healthcare providers and to the research midwifery team who undertook pregnancy outcome data collection, with allocation only revealed if medically necessitated. Once enrolled in the study, participants were given a personal study code to maintain anonymity and confidentiality during their participation. Data entry, security and storage followed Swansea University data protection policy available at https://www.swansea.ac.uk/about-us/compliance/data-protection/data-protection-policy/.

### 2.5. Physical exercise intervention programme

A 20-week supervised physical exercise programme for pregnant women was specifically designed based on i) the main antenatal exercise recommendations of the American College of Obstetricians and Gynaecologists (23); ii) evidence that a combination of cardiorespiratory and resistance exercise should be recommended for pregnant women (53–57); and iii) different exercise protocols in experimental studies (58,59). In addition, pelvic floor muscle training exercises were included in the physical exercise programme given the strong evidence that pelvic floor muscle training during pregnancy helps to prevent stress urinary incontinence (60– 62). Moreover, the physical exercise programme was designed following the Frequency, Intensity, Time, Type, Volume and Progression (FITT-VP) principles established by the American College of Sports Medicine (63). The concept of ‘regression’ was also included in the physical exercise programme -we considered it appropriate to regress some exercises and movements, especially in late pregnancy (e.g., up-right push ups against a wall instead of regular push-ups or knee push-ups). The intervention was conducted in a group, however, exercises could be individuals tailored depending on the participant’s ability or possible discomfort (e.g., lunges were substituted for standard squats if a participant felt discomfort in the symphysis pubis). The physical exercise programme was designed and delivered by the first author ORR PhD in Sport Science specialised in exercise during pregnancy. The programme was delivered at the physiotherapy gym facility at two local hospitals. Both rooms were well ventilated, with a temperature of 20º Celsius. Changing and toilet facilities were available next to each of the exercise rooms.

Participants in the INT were enrolled into the programme between 12- and 16-weeks pregnant and were expected to attend group classes up until 36 weeks pregnant. Beyond 36 weeks pregnant, participants decided whether they wanted to continue attending classes. Exercise prescription for these participants was focused on pelvic movements to enhance pelvic adaptation during labour and birth (ESM 4, 63).

Sessions were provided during the morning, afternoon, and evening from Monday to Friday, in which participants were asked to attend two or three sessions each week. All sessions each week were identical to ensure participants received the same exercise stimulus irrespective of the sessions attended. Participants were also encouraged to stay active during pregnancy in order to comply with national and international PA recommendations for pregnant women (23, 64). Participants were asked to complete a minimum of a 50% of the programme (30 exercise sessions) with a preferred attendance of at least 80% (48 exercise sessions). Attendance was monitored with adherence derived from the total number of exercise sessions and overall percentage attendance.

Exercise sessions lasted for 60 minutes and were divided into three parts: i) warm-up for 10 minutes consisting of joint movements (e.g. shoulder rotations, pelvic circles, thoracic mobility, etc), aerobic coordinative movements (e.g. double step touch, toe tipping with the addition of different arms moves), basic movement patterns (e.g. knee and hip-dominant movements) and core activation (including pelvic floor and breathing exercises); ii) 40 minutes of structured circuit training including cardiorespiratory and resistance exercises; iii) cool-down for 10 minutes, consisting of dynamic stretching of the main body structures trained during the class and pelvic movements, including pelvic tilts and circles. Example of typical classes are presented in Figure 2. Moreover, three videos have been created to bring a more graphical example of the classes (ESM 4).

**Figure 2.**
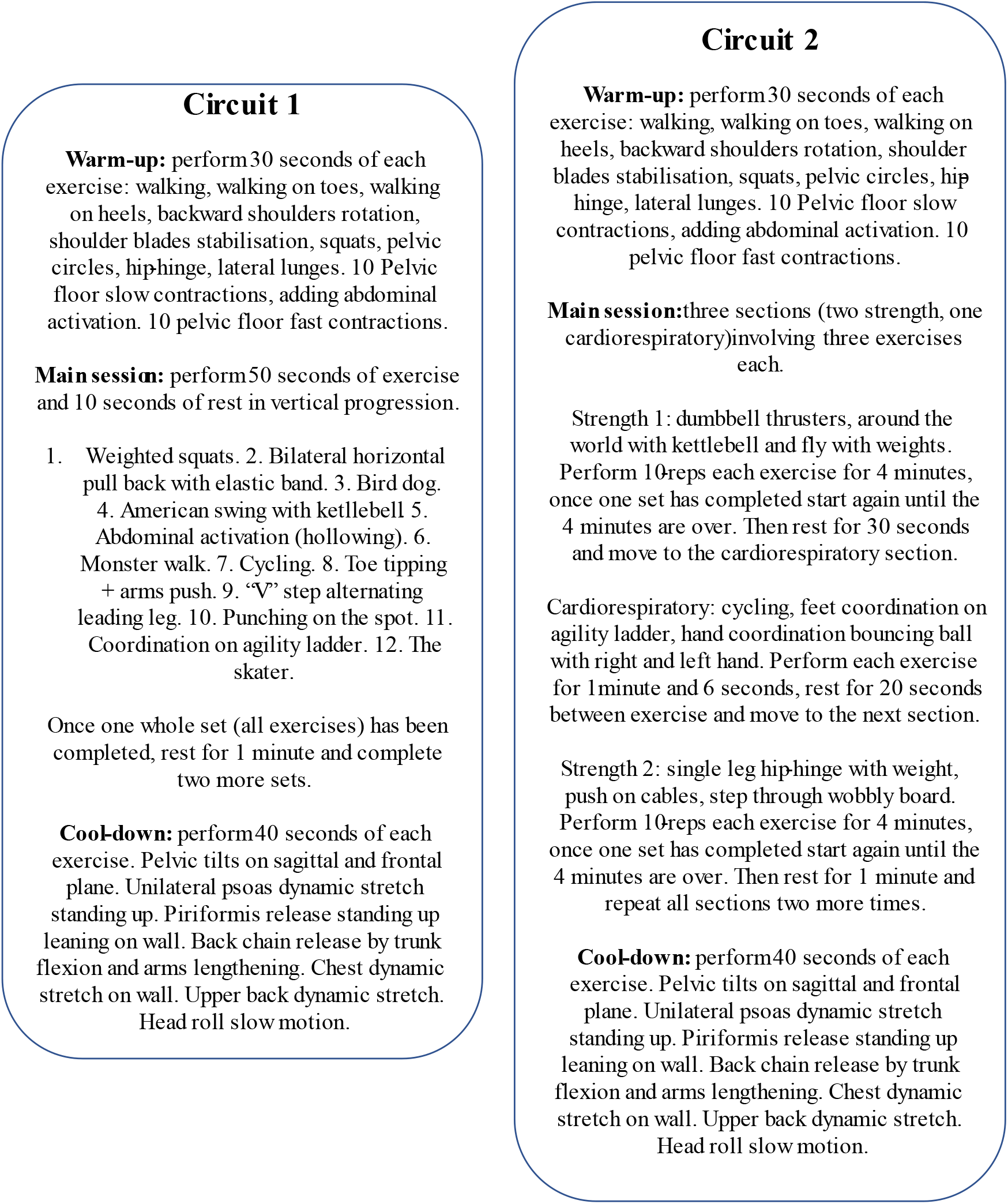
Example of the two different circuits.

The intensity of the classes was set using Brog’s rating of perceived exertion (RPE) 6 to 20 scale (66) which participants were familiarised with prior to starting the physical exercise programme. Participants‘ RPE was monitored during and after the sessions, with a summary score recorded at the end of each exercise session. Women who were sedentary prior to pregnancy were encouraged to maintain a lower intensity (11-12 on Borg’s scale) for the first four weeks (67) of the physical exercise programme with a gradual increase to moderate intensity (12-16 on Borg’s scale), while previously active women were encouraged to maintain a moderate intensity (12-16 on Borg’s scale). Regardless of each participant’s pre-pregnancy physical activity levels, the intensity was gradually increased from 16 to 20 weeks of gestation Borg’s scale 12 to 14), reaching a plateau from 20 to 36 weeks (Borg’s scale 12 to 16), and afterwards was reduced to light intensity (Borg’s scale 10 to 13). In order to individually progress with intensity levels and ensure all participants reached a moderate intensity level based on RPE, if a participant scored lower or higher than in comparison to two consecutive sessions, then the next session was progressed or regressed, as appropriate, for them. Progression was done by an increase in weight load during strength exercises, in speed in cardiorespiratory exercises. Regression was enabled by a decrease in weight load during strength exercises, decrease speed in cardiorespiratory exercises and/or the adaptation of certain movement patterns, along with the inclusion of more mobility exercises.

The exercise equipment used to deliver the programme included: two Monark exercise bikes, set of paired dumbbells (1, 2, 3 and 5kg), set of kettlebells (4, 6, 8, 10 and 12kg), wobble board, Swiss ball (65cm diameter), agility ladder, elastic bands (medium (6.80-15.80 kg) and heavy (11.34-29.50 kg) resistance), glut bands (light, medium and heavy resistance), fabric glut band medium resistance and yoga mats.

### 2.6. Study procedures

All participants were assessed on the three time-points mentioned above. A standardised experimental protocol was used for data collection, including body composition assessment, upper-body flexibility and strength assessment, and haemodynamic and cardiovascular evaluation before, during and after exercise. All physiological and anatomical variables were assessed non-invasively.

#### 2.6.1. Body composition assessment

Body mass and stature were measured at the beginning of the protocol. A Seca 803 digital scale (Seca, Hamburg, Germany) was used to measure body mass to the nearest 0.01kg and a Seca 206 wall stadiometer (Seca, Hamburg, Germany) was used to measure stature to the nearest 0.01m. Maternal body composition was assessed via skinfold thicknesses and regional body circumferences following standardised protocols from the International Society for the Advancement of Kinanthropometry (68), using a Harpenden skinfold calliper (Baty, England, UK) and ergonomic circumference measuring tapes (Seca, Hamburg, Germany; and Lufkin W606PM, Apex-tool Group, Maryland, USA). Seven skinfolds were measured (biceps, triceps, subscapular, supracristale, ilio-spinale, thigh and calf) to obtain an estimation of body fat percentage according to the equations of Paxton et al (69). Ten circumferences were measured (neck, relaxed arm, flexed-and-tensed arm, wrist, chest, waist, umbilical waist level, hips, mid-thigh and calf) to provide more information about physical changes in the prenatal and postnatal periods. Waist circumference/umbilical waist level circumference and hip circumference were used to calculate waist-to-hip ratio (70,71). Each skinfold and circumference were measured a minimum of three times, with additional measurements taken if consistency (difference between measurements greater than 0.5 cm) was not achieved within these values.

#### 2.6.2. Upper-body strength and flexibility assessment

Maximal hand-grip strength was assessed bilaterally following the protocol described elsewhere (72) using an analogue Takei 5001 hand-grip dynamometer (Takei Scientific Instruments, Japan). Briefly, participants stood still in the standard anatomical position holding the dynamometer (arms straight down by the trunk sides). At the researche’s signal they applied a maximal grip squeeze without flexing the elbow or pushing the arm against the trunk (if either of these deviations occurred the attempt was considered invalid and repeated). Each hand was tested alternately, twice for each hand, with the maximum score achieved on each hand used in subsequent analyses.

Upper-body flexibility was measured using the back-scratch test (73) which assesses the range of motion of the shoulder by measuring the distance between, or the overlap of the middle fingers behind the back using a Lufkin tape measure. Two attempts with alternate positioning of the arms (right arm up and left arm down, and vice versa) were performed and then repeated. The best score (minimum distance from right middle finger to left middle finger or maximum overlap between those fingers) for each side was noted for subsequent analysis.

#### 2.6.3. Haemodynamic and cardiovascular assessment and exercise protocol

Maternal haemodynamic and cardiovascular assessment were performed continuously throughout the exercise protocol, which was three stages as described below. ECG recordings were obtained using a Holter monitor (Lifecard Digital system; Spacelabs Medical Ltd., UK) with a sampling frequency of 1024Hz. Haemodynamic variables were obtained via the Task Force Haemodynamic Monitor (Task Force Monitor, CNSystems Medizintechnik GMBH, Austria). Both of these methods have been used with pregnant women previously, showing their reproducibility (14, 35, 73).

FHR was continuously monitored before, during and after the exercise bout via cardiotocography (CTG; Huntleigh, Sonicaid Foetal Monitor – Team 3 series, USA) during time-point 2 only. FHR was recorded electronically using software (Sonicaid Centrale Hospital System, Huntleigh, USA) which allowed archiving of the CTG trace and CTG data extraction for analysis.

##### First stage of the exercise test

Maternal haemodynamic and cardiovascular measurements were initially performed throughout a five-minute period of seated rest. This resting period was used to calculate target HR zones for the subsequent exercise test of between 40% to 60% of heart rate reserve (HRR) for individuals using the Karvonen formula (51,55):

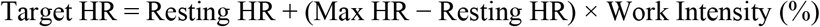

Where resting HR was calculated via the arithmetic average of maternal resting HR observed every 30 seconds during the seated resting stage, max HR was calculated using the equation developed by Tanaka, et al. (75) for healthy individuals, and work intensity was either 0.4 or 0.6 to calculate 40% or 60% of HRR_(maxHR-restingHR)_, respectively.

FHR was recorded during this stage but only during the assessment time-point 2. This was done over a period of 20 minutes while the participant remained seated in order to obtain two 10-minute sections for subsequent FHR analysis, according to FIGO guidelines (76,77). Maternal HR was assessed for this period as recommended by the International Federation of Gynaecologists and Obstetrics (FIGO) (78), however only the last five minutes of maternal ECG and haemodynamic recordings were used for analysis to maintain consistency with the other two assessment points.

##### Second stage of exercise test

Maternal cardiovascular measurements were performed continuously throughout 10-minutes of stationary upright bicycle exercise (Monark 824E, Monark, Sweden) at a moderate intensity (40-60% HRR; 50, 54). This stage began with a five-minute warm-up at a cadence of 50 revolutions per minute (rpm) at a 50W resistance. Next, the participant cycled at 60rpm, cycling speed was increased by 5 rpm every two-minutes to elicit a corresponding 5W increase until the participant’s HR reached the lower threshold of her target HR zone (40% HRR). Cadence was then maintained for the remainder of the test.

Foetal CTG was measured throughout this stage at the assessment time-point 2.

##### Third stage of exercise test

Maternal cardiovascular measurements were assessed continuously throughout a 20-minute recovery period. Once the 10-minute cycling was completed, one-minute of active recovery was performed on the cycle ergometer at 50rpm. Subsequently, participants completed a 19-minute passive, seated, recovery period. Foetal CTG was measured throughout this stage at the assessment time-point 2.

#### 2.6.4. Assessment of habitual physical activity levels

Estimations of each participant’s habitual PA levels were obtained subjectively using two self-administered questionnaires, the Modifiable Activity Questionnaire (79,80) and the Pregnancy Physical Activity Questionnaire (81). Data obtained from these questionnaires provided information on participants‘ PA habits during the three months prior to conception (retrospectively reported) to the three months after birth.

Device-based PA levels were recorded from a random sub-group of participants from both the INT (n=10) and COMP (n=10). Participants were asked to wear a wrist-worn accelerometer (Actigraph GT9X – Actigraph Corp., Pensacola, USA) on the non-dominant wrist for a seven consecutive days period following each of the three-assessment time-point. Data from these devices provided quantified information on time spent at light, moderate and high PA, sedentary time, and sleep patterns.

### 2.7. Primary outcome measures

The primary outcome measure of the study is a multi-variable assessment of maternal cardiovascular function (based on SV, HR and HRV). The outcome measures will be defined in terms of ‘change values’. Two periods of ‘change’ will be considered: i) ‘antenatal change’ from mid to late pregnancy and ii) ‘postnatal change’ from late pregnancy to 12-16 weeks postpartum, enabling a longitudinal assessment of the main cardiovascular and haemodynamic changes during pregnancy and the early postnatal period.

Maternal cardiovascular variables were derived from the Holter ECG monitoring (Lifecard Digital system; Spacelabs Medical Ltd., UK) and from the Task Force Haemodynamic Monitor (Task Force Monitor, CNSystems Medizintechnik GMBH, Austria). The main variables extracted from Holter ECG recordings were beat-to-beat cardiac interval (R-R interval), RMSSD, SDRR and the QT interval (onset of Q wave to end of T wave). The main variables obtained from the Task Force Monitor were SV, HR, systolic and diastolic BP, 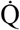 and cardiac index (the cardiac output divided by body surface area), left ventricular ejection time and EDI, each of which were recorded on a beat-to-beat basis.

### 2.8. Secondary outcome measures

The secondary outcome measures from the study relate to FHR, maternal health-related fitness (muscular strength, flexibility and body composition), and to a series of indices derived from questionnaires that sought to evaluate whether prenatal exercise influences perceived quality-of-life, urinary incontinence and maternal and offspring sleeping patterns.

FHR obtained from CTG recordings will be analysed to evaluate the effect of the supervised physical exercise programme on the FHR response to an acute bout of maternal exercise (10-minute cycling as described above).

Another set of questionnaires was completed by all participants to obtain information on the possible influence of prenatal exercise on quality-of-life scores, urinary incontinence, maternal sleeping patterns and offspring sleeping patterns. Questionnaires were completed three times coinciding with the scheduled assessment dates.

The QoL-Grav questionnaire (82) was chosen to evaluate quality of life in pregnant women and the influence thereon of the physical exercise programme. It has been validated and it is more sensitive to the specific psychological and physical changes occurring in the life of a pregnant woman.

The International Consultation of Incontinence Questionnaire-Urinary Incontinence (83) was administered to participants to investigate symptoms of urinary incontinence, and the influence thereon of the physical exercise programme. This questionnaire has been widely used within the pregnant population (60,84–86).

The Pittsburgh Sleep Quality Index (87) and the Insomnia Severity Index (88) were utilised to study sleeping patterns in pregnancy and to examine whether exercise influences sleep quality in pregnancy in a similar way to that observed in the non-pregnant population (89).

Offspring sleep patterns were studied via the Brief Infant Sleep Quality Questionnaire (90). This will be used to investigate whether maternal PA habits during and after pregnancy influence offspring sleeping patterns. Furthermore, information from this questionnaire can be analysed in conjunction with maternal sleep patterns and quality of life, as suggested in Pires et al. (91).

### 2.9. Statistical analysis

A series of analyses will be carried out to determine the effect of the exercise intervention on the primary and secondary outcomes of the study. Statistical analysis will be performed on a per-protocol and intention-to-treat basis.

Normality and homogeneity of the data will be assessed. If confirmed, independent sample t-test analyses will be carried out for the demographic description of the sample and for comparison of pregnancy outcome between groups. To assess the intervention effect on primary outcome variables and FHR, general linear models will be performed and reported as between-group mean difference. Correlations will be performed to study the relationship between the time spent in light, moderate and high intensity PA at the three-assessment time-point on pregnancy outcome, sleep quality, new-born sleep patterns, QoL, urinary incontinence, body composition, hand-grip strength, and upper body flexibility.

When normality and homogeneity of the data are not confirmed, alternative non-parametric analyses will be performed.

### 2.10. Data dissemination

Only the research team will have access to the final trial data set. Partial publication of the final data set can be shared upon request.

Data will be disseminated via open access publication in relevant scientific journals, and national and international conferences. Data will be shared with participants and health care providers in a face-to-face event.

## 3. Discussion

Within this paper we have described the protocol of the PE-CAMP study to evaluate the impact of an antenatal supervised physical exercise programme on non-invasive estimates of cardiac and haemodynamic function in the mother and cardiac function in the foetus.

This study will enable: i) characterisation of some key cardiac and haemodynamic biomarkers in the pregnant woman and her foetus; ii) evaluation of the influence of antenatal exercise on these biomarkers; and iii) evaluation of changes in maternal biomarkers during the early postnatal period.

One of the most novel features of the study is the thorough and standardised exercise protocol which replicates the minimum duration of PA (10 minute bouts) recommended to pregnant women by the UK Chief Medical Officer (65). The first version of these guidelines released in 2017 state that a minimum of 150 minutes of moderate PA should be achieved throughout the week, accumulated in at least 10-minute bouts, to obtain health benefits. We therefore replicated an acute exercise bout of 10 minutes to estimate maternal and foetal responses to an achievable and nationally recommended PA prescription. The exercise intervention was designed by a sports scientist specialised in prenatal exercise and draws on relevant research in the field (50, 91, 92). This gives a strong foundation to our exercise intervention which may elicit beneficial adaptations in the maternal cardiovascular system, since it follows official recommendations and evidence-based interventions that have already demonstrated benefits in pregnant women.

Although it is necessary and informative to continue investigating the effects of exercise on maternal cardiac and haemodynamic responses using specific laboratory-based tests, it is also critical to evaluate these influences during physical activities that are more achievable and realistic for pregnant women during their daily lives. The PE-CAMP study will provide data on the cardiac and haemodynamic responses to a typical acute bout of exercise, that could help inform future decisions and policy on maternal exercise prescription made by maternity healthcare providers and exercise professionals. It is vital for healthcare providers, researchers and exercise professionals working with the pregnant population to fully understand such prenatal responses to exercise to individualise the exercise prescription and to design safe and effective exercise programmes to be performed by pregnant women.

One of the greatest strengths of the PE-CAMP Study is the supervised exercise intervention, which enables the research them to objectively quantify participants’ exercise levels and exercise prescription. This provides confidence that any differences in the biomarkers observed in the INT and COMP groups can be attributed to the physical exercise programme. Moreover, habitual PA and exercise levels were assessed both using devices (for a sub-group of the sample, using accelerometry) and subjectively (using questionnaires), facilitating pre-conception and pregnancy comparisons.

Although the PE-CAMP has several strengths, there are limitations which warrant acknowledging. Specifically, women in the COMP will be advised to follow usual advice provided by their main healthcare providers (i.e. maintaining a healthy lifestyle) and may therefore participate in other exercise programmes or engage in sufficient PA levels. As such women in the COMP who achieve or exceed the minimum PA recommendation for pregnant women (150 min• week^-1^), assessed via questionnaires, will be excluded from our main statistical analysis, since this might confound results from participants in the INT. Data from these participants might be used for further analysis to observe the effect of non-supervised exercise on the maternal/foetal cardiac response to acute and chronic exercise.

FHR assessed via CTG may be subject to signal loss due to foetal and maternal movements and the use of a paper trace which reduces the subsequent depth of post-prior analyses. However, the specific CTG equipment used in our study allows us to transfer FHR digitally for further analysis, mitigating these issues. Moreover, the assessment protocol in this study involves a bout of exercise performed on a cycle-ergometer, which should minimise maternal movement and help prevent signal artifact in the devices used (maternal ECG, ICG and FHR).

## Supporting information

ESM1: SPIRIT Checklish

ESM2: CERT Checklist

ESM3: Consent Form

ESM4: Link to intervention videos

## Data Availability

Data produced in the present study are available upon reasonable request to the authors.

https://www.youtube.com/watch?v=QTbuO-ivT_U

https://www.youtube.com/watch?v=_6sPOV2e0GU

https://www.youtube.com/watch?v=v8-bJWsC1ac

## Acknowledgments

This project has been fully funded under the fellowship of ORR, which was approved by the Sêr Cymru 2 Fellowship Programme, co-funded by the Welsh Government, the European Regional Development Fund and Swansea University. COFUND, Marie Sklodowska-Curie Actions (grant agreement number 663830).

The research team would like to acknowledge the contribution of the research midwifery team and the staff team at the antenatal clinics where the study was conducted. Without your help and support this study would have not been possible.

## Trial sponsor information

Paola Griffiths, Research Governance Officer, Swansea University.

Tel: 01792 606060

