## Supplementary material for "The Physical Exercise and Cardiovascular Adaptation Monitoring in Pregnancy (PE-CAMP) Randomised Controlled Trial Protocol": ESM2: CERT Checklist

### CERT ✓ Consensus on Exercise Reporting Template

#### A Checklist for what to include when reporting exercise programs

| Section/Topic | Item # | Checklist item | Location ** | |
|---|---|---|---|---|
| | | | Primary paper (page, table, appendix) | † Other (paper or protocol, website (URL) |
| **WHAT: materials** | 1 | Detailed description of the type of exercise equipment (e.g. weights, exercise equipment such as machines, treadmill, bicycle ergometer etc) | 15 | |
| **WHO: provider** | 2 | Detailed description of the qualifications, teaching/supervising expertise, and/or training undertaken by the exercise instructor | 12 | |
| **HOW: delivery** | 3 | Describe whether exercises are performed individually or in a group | 12 | |
| | 4 | Describe whether exercises are supervised or unsupervised and how they are delivered | 12 | |
| | 5 | Detailed description of how adherence to exercise is measured and reported | 13 | |
| | 6 | Detailed description of motivation strategies | N/A | |
| | 7a | Detailed description of the decision rule(s) for determining exercise progression | 15 | |
| | 7b | Detailed description of how the exercise program was progressed | 15 | |
| | 8 | Detailed description of each exercise to enable replication (e.g. photographs, illustrations , video etc) | 14 & ESM 2 | |
| | 9 | Detailed description of any home program component (e.g. other exercises, stretching etc) | N/A | |
| | 10 | Describe whether there are any non-exercise components (e.g. education, cognitive behavioural therapy, massage etc) | 8 & 13 | |
| | 11 | Describe the type and number of adverse events that occurred during exercise | N/A | |

| | | | |
|---|---|---|---|
| **WHERE: location** | 12 | Describe the setting in which the exercises are performed | 12 |
| **WHEN, HOW MUCH: dosage** | 13 | Detailed description of the exercise intervention including, but not limited to, number of exercise repetitions/sets/sessions, session duration, intervention/program duration etc | 12 & 14 |
| **TAILORING: what, how** | 14a | Describe whether the exercises are generic (one size fits all) or tailored whether tailored to the individual. | 12 |
| | 14b | Detailed description of how exercises are tailored to the individual | 12 |
| | 15 | Describe the decision rule for determining the starting level at which people commence an exercise program (such as beginner, intermediate, advanced etc) | 11 & 15 |
| **HOW WELL: planned, actual** | 16a | Describe how adherence or fidelity to the exercise intervention is assessed/measured | 13 |
| | 16b | Describe the extent to which the intervention was delivered as planned | 15 |

***It is recommended that this checklist is used in conjunction with the Explanation and Elaboration Statement which is a guide each item in the CERT Checklist**

The CERT Checklist is designed for reporting details of an exercise intervention. The CERT Checklist should be used in conjunction with a reporting checklist appropriate for the study type e.g. the CONSORT Statement (www.consort-statement.org) for randomised controlled trials, the SPIRIT Statement (www.spirit-statement.org) for a clinical trial protocol. For further guidance regarding reporting guidelines please consult the EQUATOR network (www.equator-network.org)

** Authors – please use N/A if an item is not applicable Reviewers – please use ”?” if information is not provided or not/insufficiently reported

† If the information is not provided in the primary paper that is under consideration, please provide details of where this information is available e.g. in a published protocol, published papers (provide citation details) or on a website (provide the URL).
