## Supplementary material for "The Physical Exercise and Cardiovascular Adaptation Monitoring in Pregnancy (PE-CAMP) Randomised Controlled Trial Protocol": ESM3: Consent Form

ESM 3: Informed consent form. PE-CAMP Study, Consent Form, v5 (13/02/2019), IRAS ref: 239704

**CONSENT FORM (Version 5)**

**IRAS ref: 239704**

Participant Identification number

N.B. Three copies will be made for
(1) Participant
(2) Researcher
(3) Hospital notes

**Title of Project:** **Physical exercise and cardiovascular adaptation monitoring in pregnancy (PE-CAMP)**

**Name of Researchers:** [redacted]

**Contact Telephone Number:** [redacted]

**Please Initial Box**

1. I confirm that I have read and understood the information sheet dated 13/02/2019 (Version 6) for the above study. I have had the opportunity to consider the information and to ask questions, and these have been answered satisfactorily.

2. I understand that my participation is voluntary and that I am free to withdraw at any time, without giving any reason, without my medical care or legal rights being affected.

3. I understand that relevant sections of my medical notes and data collected during the study may be looked by individuals directly involved in the study, from the University or from the NHS Health Board, where it is relevant to my taking part in this research. I give permission for these individuals to have access to my records.

4. I agree to wear the physical activity monitor (Actigraph) for 7 days. I understand that I am free to stop using it at any time, without giving any reason, without my medical care or legal rights being affected.

5. I agree to my GP being informed of my participation in the study

6. I agree to take part in the above study. My results may be used in an anonymous fashion in a future publication in a medical journal.

7. I understand that I will not be paid for taking part in this study but I may be reimbursed for reasonable travelling costs to Singleton hospital.

8. I consent to be contacted by Sarah Fox (research Midwife) to discuss my experience of participating in this research **once my participation**

**in this study has ended.**

| | | |
|---|---|---|
| Name of Participant | Date | Signature |
| Name of person taking consent | Date | Signature |
