## Supplementary material for "The Physical Exercise and Cardiovascular Adaptation Monitoring in Pregnancy (PE-CAMP) Randomised Controlled Trial Protocol": ESM4: Link to intervention videos

**ESM 4: Links to physical exercise programme videos.**

Physical exercise programme: Circuit 1.

Physical exercise programme: Circuit 2.

Physical exercise programme: Movements that can be applied during birth.
